# Excluded causal variants produce false discoveries in genetic fine-mapping

**DOI:** 10.64898/2026.11.09.26367644

**Authors:** Josh Weinstock

**Affiliations:** Emory University, Department of Human Genetics

## Abstract

Statistical fine-mapping seeks to distinguish causal genetic variants from correlated variants in linkage disequilibrium (LD). Although fine-mapping algorithms perform well in simulations, they are often applied in settings that violate their assumptions. In particular, genome-wide association study (GWAS) and quantitative trait locus (QTL) workflows routinely omit variants with minor allele frequency (MAf) below 1% and most insertions and deletions (indels), excluding many causal variants before fine-mapping begins. To assess this empirically, we compared filtered and unfiltered fine-mapping at 499 All of Us loci. Twenty percent of high-confidence calls after a 1% MAf filter, and 32% when indels were also omitted, lacked support in the unfiltered analysis. Errors were particularly enriched when tested variants tagged an omitted variant, so fine-mapping misassigned its signal to a proxy variant. At *RBO*, omitting the frameshift that defines blood group O gave near-certain posterior inclusion probabilities (PIPs) to linked single-nucleotide polymorphisms (SNPs). To address this disconnect, we developed SuSiE-OUT, which enables fine-mapping in scenarios with possibly omitted causal variants. Modeling infinitesimal effects did not reliably prevent these errors, whereas imputing omitted variants or modeling them with our SuSiE-OUT method prevented most errors. Overall, we find that common analysis workflows substantially increase false discoveries in fine-mapping analyses, and we demonstrate two approaches to mitigate this challenge.

## Introduction

A common mission of human genetics is the discovery and characterization of genetic variation that causes phenotype variation. In the past two decades, genome-wide association studies (GWAS) have mapped thousands of trait-associated regions. However, nearby alleles are correlated (linkage disequilibrium, LD), so the variant with the strongest association is often not the causal variant [1, 2, 3, 4, 5]. Statistical fine-mapping addresses this problem. It assigns each variant a posterior inclusion probability (PIP), the probability that the variant contributes to the association, and it groups variants into credible sets designed to contain a causal variant with a stated probability [6, 7, 8, 9, 10]. functional annotations and cross-population LD can further reduce the number of candidate variants [11, 12, 13, 14]. Downstream functional analyses routinely use high-PIP variants to link associations to genes and mechanisms and to choose variants for experimental follow-up [15, 16, 17, 18, 19, 20, 21].

Standard fine-mapping models assign posterior probability only among the variants included in the analysis. They therefore implicitly assume that every causal effect can be represented by those variants [8, 9, 22]. Common analysis workflows routinely violate this assumption, because they omit variants before fine-mapping. Minor allele frequency (MAf) thresholds, most often 1%, omit rare variants which may be causal [23, 24, 25]. Many analyses are also restricted to single-nucleotide polymorphisms (SNPs), because LD references, imputation panels and public fine-mapping releases exclude insertions and deletions (indels) [26, 27, 28, 29, 30]. These filters preferentially omit variants that are likely to have large effects. Purifying and stabilizing selection keep larger-effect alleles rare [31, 32, 33, 34], and indels include frameshift and in-frame coding changes that are among the most consequential variants in the genome [35, 26].

Omitting a causal variant does not remove its association signal. When the omitted variant is correlated with the variants retained for analysis, its conditional effect is implicitly included in the marginal association statistics of tested variants. Sparse fine-mapping must then explain the marginal associations of tested variants, and it can give near-certain PIP to a correlated noncausal variant, which we call a *pvoxy* [6, 5, 22] (fig. 1). The result is a confident but incorrect causal interpretation with two consequences: functional follow-up targets a variant with no effect, and, when the omitted variant is a coding change, the analysis misses the most direct mechanistic explanation for the signal. We expected the frequency of such errors to depend on tagging, the extent to which tested variants associate with an omitted variant. A weakly tagged variant leaves little association among tested variants to misassign. In contrast, a well-tagged variant can produce a strong proxy. Summary-statistic imputation can recover well-tagged variants before fine-mapping [36, 37]. However, summary-statistic imputation is not a standard step, and previous studies have not measured how often filtering produces proxy calls.

**Fig. 1.**
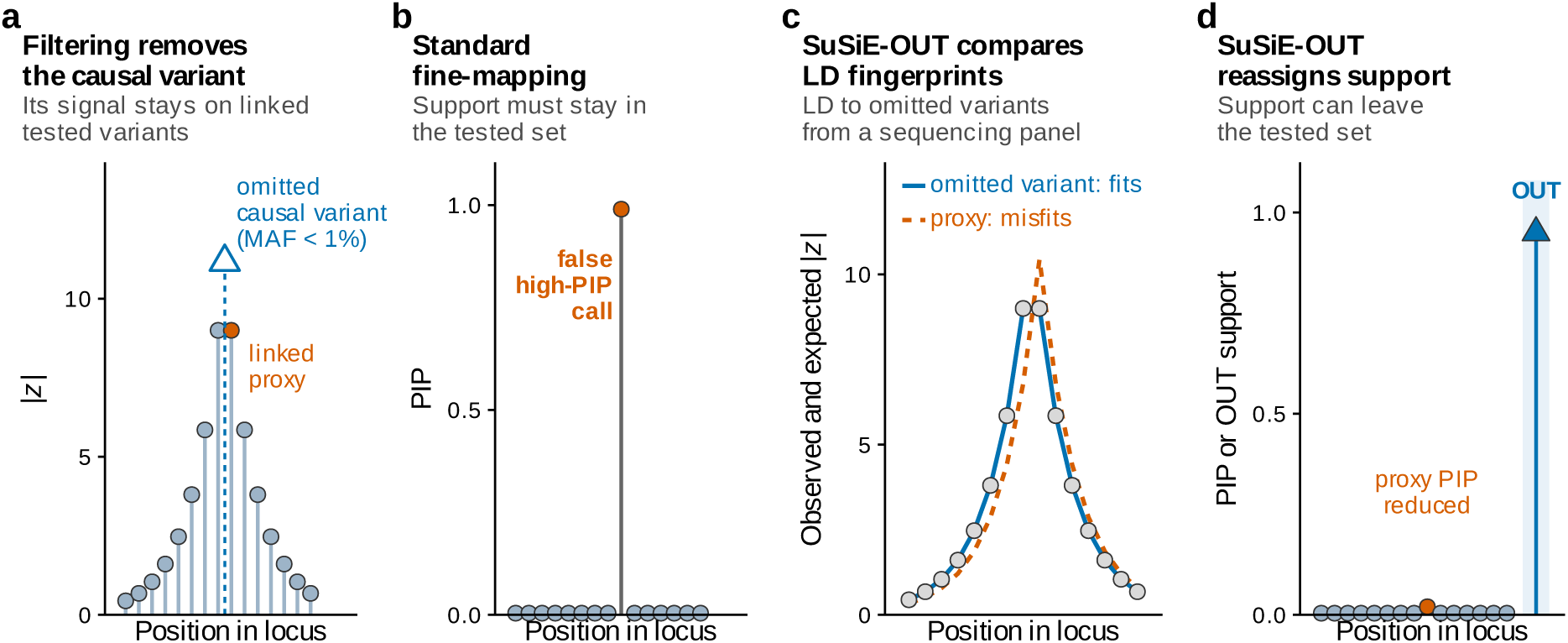
Filtering a tagged causal variant creates false high-PIP calls. Schematic of a single locus. **a,** A frequency or variant-type filter omits the causal variant (open triangle), but linked tested variants (circles) still show its association. **b,** Ordinary fine-mapping must assign that association to a tested variant and can give a correlated proxy (orange) near-certain posterior inclusion probability (PIP). **c,** SuSiE-OUT compares the observed *z* pattern (points) with the pattern expected from each candidate’s LD fingerprint (its correlations with the tested variants). Candidates include omitted variants whose LD is known from a sequencing panel. **d,** SuSiE-OUT can therefore assign the effect to the omitted variant (an OUT state), and the proxy’s PIP falls.

Here we ask three questions about two routine analysis filters: a 1% MAf threshold and a SNP-only filter. first, how often does filtering create high-PIP calls that an unfiltered analysis does not support? Because sequencing provides association statistics for every variant, we could answer this by comparing fine-mapping with and without the filters at 499 All of Us loci [38, 39], and we extended the comparison to public UK Biobank (UKB) fine-mapping [40, 41, 30]. Second, why do these errors arise, and when are these errors most severe? We find that they concentrate where tested variants tag the omitted variant. Third, can the errors be prevented when association statistics for the omitted variants are unavailable? To assess this, we built an extension of SuSiE called SuSiE-OUT that explicitly models the probability a causal variant was omitted. Rather than forcing every association signal onto a tested variant, SuSiE-OUT allows a signal to originate from an omitted variant. SuSiE-OUT only needs the LD between tested and omitted variants, which can be estimated in an appropriate LD reference panel. Overall, we quantify the consequences of common analysis procedures on fine-mapping accuracy, and we develop measures to mitigate common failure modes.

## Results

### Filtering changes fine-mapping results

To assess the impact of common filtering procedures, we analyzed 499 independent genome-wide-significant All of Us loci [39]: 367 for height (*n* = 275,875), 101 for platelet count (*n* = 141,410) and 31 for low-density lipoprotein cholesterol (LDL-C; *n* = 137,377), three traits with well-characterized genetic architectures [42, 43, 44]. All analyses used the European-ancestry group in which we discovered the loci and estimated LD (Methods). At each locus we applied two filters that reflect common practice: the *MRF filtev*, which keeps variants with MAf of at least 1% and so omits rare variants, and the *SMK-oely filtev*, which keeps only SNPs with MAf of at least 1% and so also omits every indel. We call the variants that pass a filter *tested* variants and the variants that it excludes *omitted* variants. We fit SuSiE [9] to the tested variants (*masked-SuSiE*) and to all variants that passed quality control (*uefilteved-SuSiE*). All fits allowed at most ten effects and used the same participants, association statistics and in-sample LD. We used the whole-exome sequencing (WES) derived genotypes; although this omits non-coding variation, where most trait-associated variants lie [45, 18], it avoids very large, near-singular genome-wide LD matrices, which can require regularization that itself alters fine-mapping results [5, 22]; we chose not to conflate these effects with those of variant filtering. Differences between masked-SuSiE and unfiltered-SuSiE therefore reflect only the variant filters.

We used unfiltered-SuSiE as a silver standard, and we refer to its PIP as the *vefeveece KIK*. We define a *call* as a variant with PIP of at least 0.90. A call is *uesuppovted* if its reference PIP is below 0.90, and an unsupported call is also a *false discovevy* if its reference PIP is below 0.10 (Table 1). In simulations, where causal status is known, a false discovery is a call on a noncausal variant.

**Table 1.**
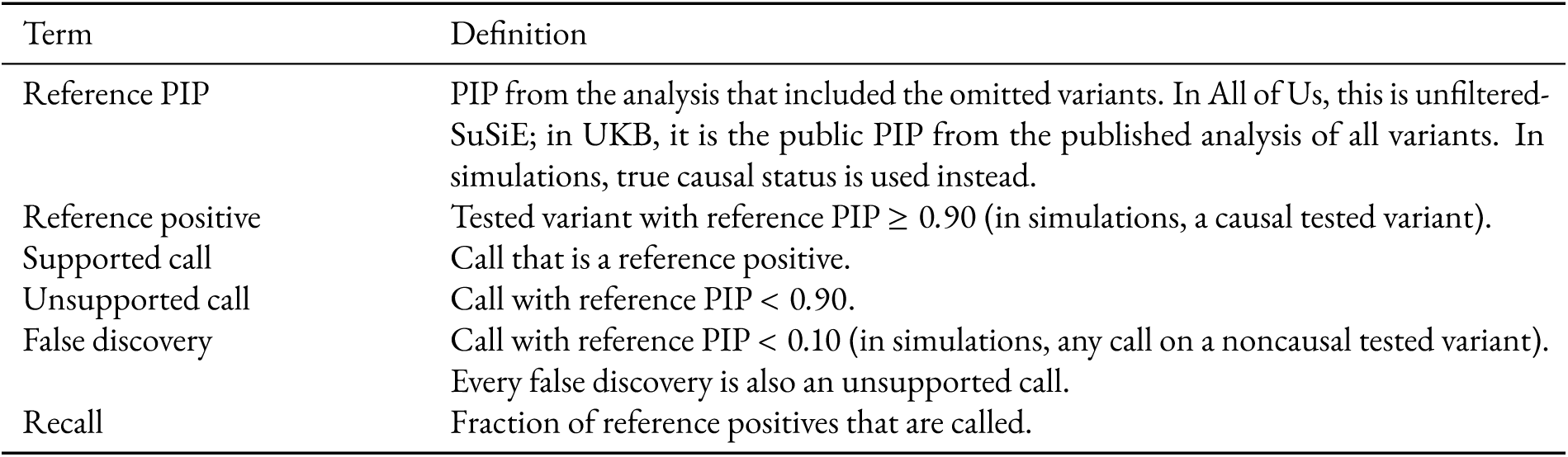
Evaluation terms. A call is any variant with PIP of at least 0.90 under the method being evaluated.

| Term | Definition |
|---|---|
| Reference PIP | PIP from the analysis that included the omitted variants. In All of Us, this is unfiltered-SuSiE; in UKB, it is the public PIP from the published analysis of all variants. In simulations, true causal status is used instead. |
| Reference positive | Tested variant with reference PIP ≥ 0.90 (in simulations, a causal tested variant). |
| Supported call | Call that is a reference positive. |
| Unsupported call | Call with reference PIP < 0.90. |
| False discovery | Call with reference PIP < 0.10 (in simulations, any call on a noncausal tested variant). Every false discovery is also an unsupported call. |
| Recall | Fraction of reference positives that are called. |

Both filters produced many unsupported calls (fig. 2a). With the MAffilter, 133 of 659 calls (20%) were unsupported and 20 were false discoveries. With the SNP-only filter, 197 of 614 calls (32%) were unsupported and 52 were false discoveries. Thus, many variants that a standard workflow would report as likely causal were not supported once the omitted variants were included.

**Fig. 2.**
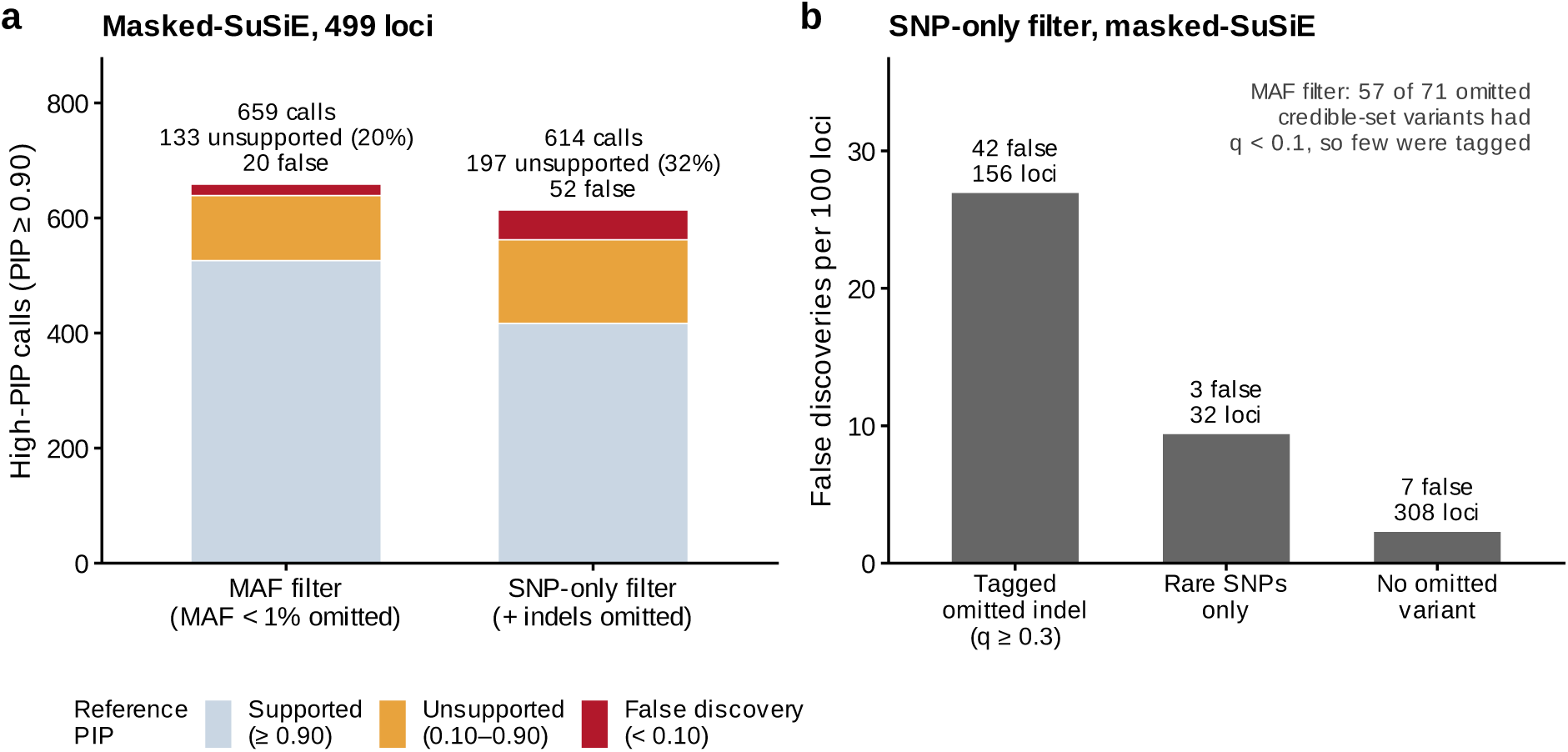
Common analysis workflows result in elevated false discoveries. Fine-mapping analysis of 499 loci for height, platelet count and LDL-C in All of Us. Each filter is scored against its own unfiltered-SuSiE fit, which includes the omitted variants. **a,** Calls (PIP 0.90) from masked-SuSiE, grouped by reference PIP: supported ( 0.90), unsupported but not a false discovery (0.10 to *<* 0.90) or false discovery (*<* 0.10). Left, MAf filter (variants with minor allele frequency below 1% omitted); right, SNP-only filter (variants with MAf below 1% and all indels omitted). **b,** SNP-only filter: masked-SuSiE false discoveries per 100 loci, by the type of omitted variant in an unfiltered-SuSiE 95% credible set: an indel tagged by the tested SNPs (tagging *q* 0.3; 156 loci), rare SNPs only (32 loci) or no omitted variant (308 loci). Three loci whose omitted indel had *q <* 0.3 are not shown. Under the MAf filter, 57 of 71 omitted credible-set variants had *q <* 0.1 (median 0.025).

### Errors concentrate where the omitted variant is tagged

Whether filtering produced a false discovery depended on how well the included variants tagged the omitted variant. Rare variants omitted by the MAf filter were poorly tagged, so the tested variants carried little of their association and few proxies arose: 85% (113/133) of unsupported calls under this filter had intermediate reference PIPs (0.10–0.90), and only 15% (20/133) were false discoveries. In contrast, indels omitted by the SNP-only filter were often common and well tagged.

We measured tagging by *q*, the fraction of an omitted variant’s association that is predicted by the tested variants; *q* equals the imputation quality of summary-statistic imputation [37] (Methods). We call an omitted variant *tagged* if *q* ≥ 0.3 and *uetagged* if *q <* 0.1. Under the MAf filter, 80% (57/71) of omitted variants in unfiltered-SuSiE 95% credible sets were untagged. Under the SNP-only filter, 98% (156/159) of loci with an omitted indel in an unfiltered credible set had indels that were tagged by SNPs. false discoveries concentrated at these 156 loci, which had 81% (42/52) of all false discoveries under this filter, or 27 per 100 loci. The 32 loci at which only rare SNPs were omitted had 9 per 100 loci (3 false discoveries), and the 308 loci with no omitted credible-set variant had 2 per 100 loci (7; fig. 2b). Calls had a mean PIP of 0.99, so if PIPs were calibrated about 8 of the 614 calls under the SNP-only filter would be noncausal. Masked-SuSiE instead made 52 false discoveries, 6.4 times this expectation, and 20 under the MAf filter (2.5 times). Omitting rare SNPs, and particularly omitting indels, therefore inflates false discoveries well beyond the rate implied by the PIPs.

To more carefully understand this phenomenon, we used simulations to study the effects of excluding causal variants at various filters. We simulated 600 loci at which MAf, indel, imputation-quality or reference-panel filters omitted some, all or none of the causal variants, under neutral, stabilizing and directional selection [32, 34] (Methods). Masked-SuSiE made at least one false discovery at more loci as the omitted causal variant became better tagged. Among loci with an omitted causal variant, this rate was 17% when the variant’s maximum *r*^2^ with a tested variant was below 0.2, 43% at 0.2–0.5 and 57% at 0.5–0.8 (fig. 3a; Supplementary Table 1). These results are consistent with tagging, rather than omission alone, producing false discoveries.

**Fig. 3.**
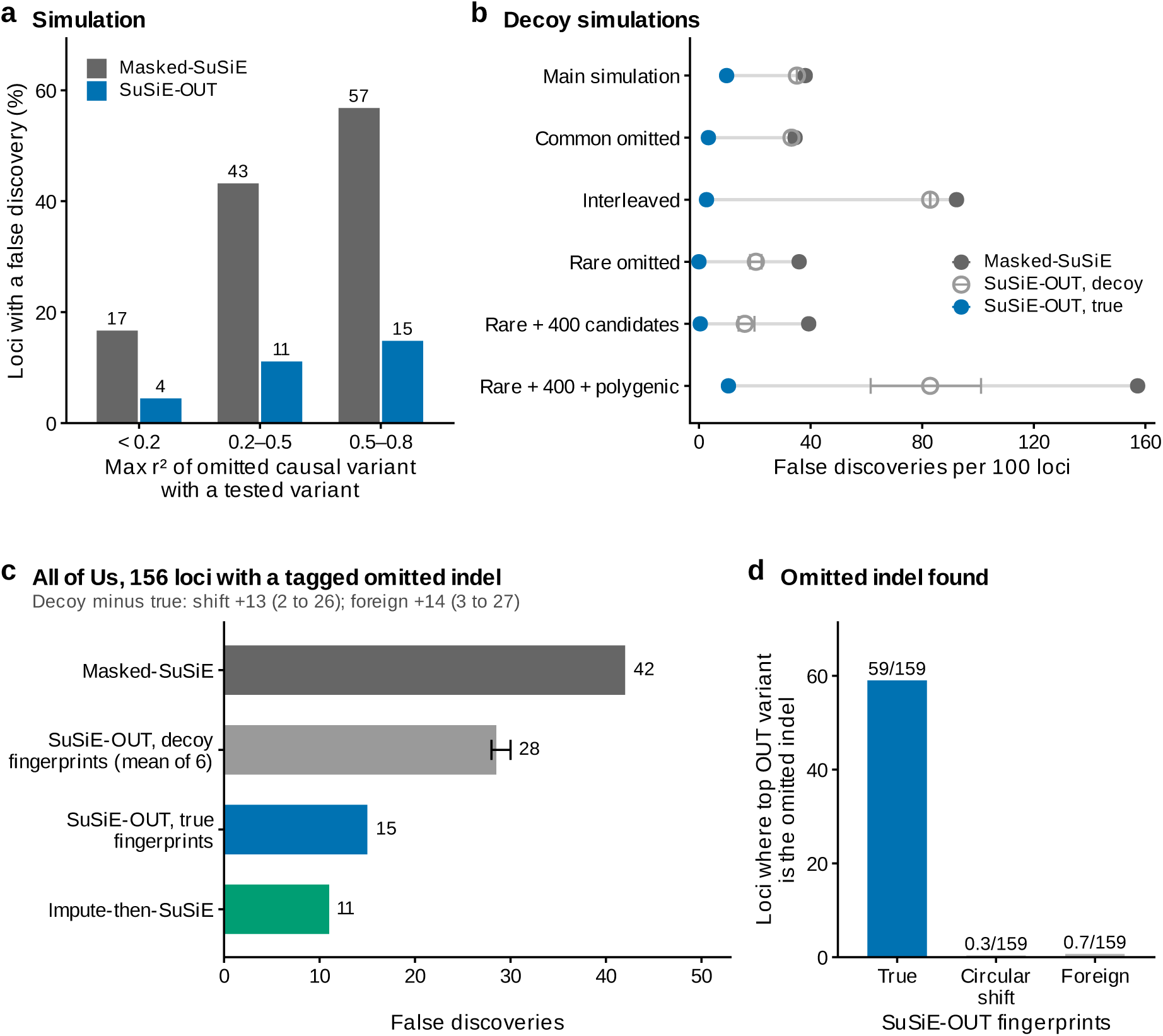
Errors increase with tagging of the omitted variant, and using its true LD fingerprint reduces them. a, Main simulation: loci with an omitted causal variant and at least one false discovery, by the maximum *r*^2^ between the omitted causal variant and any tested variant (90, 81 and 81 loci). Gray, masked-SuSiE; blue, SuSiE-OUT. **b,** Decoy simulations: false discoveries per 100 loci with an omitted causal variant, for masked-SuSiE (filled gray), SuSiE-OUT with decoy fingerprints (open symbols; mean across decoy types, with range) and SuSiE-OUT with true fingerprints (blue). Decoy types were circular shift, permutation and, in all conditions except the main simulation, foreign fingerprints from another locus. Each condition adds, alone or in combination, common omitted variants, omitted variants interleaved with tested ones, rare omitted variants, 400 OUT candidates and polygenic background; 206–252 loci each. **c,** All of Us, SNP-only filter, 156 loci where a tagged omitted indel (*q* 0.3) is in an unfiltered-SuSiE credible set: false discoveries for masked-SuSiE, SuSiE-OUT with decoy fingerprints (mean of three circular-shift and three foreign decoy sets; bar, range), SuSiE-OUT with true fingerprints and impute-then-SuSiE. Paired decoy-minus-true differences and 95% locus-bootstrap intervals (2,000 resamples) are shown above the panel. **d,** Of the 159 loci with an omitted credible-set indel, the number at which SuSiE-OUT gave its highest OUT PIP to that indel, with true or decoy fingerprints (decoys, mean of three decoy sets).

### Imputation or SuSiE-OUT corrects the errors; SuSiE-inf does not

Because these errors arise when omitted variants leave a fingerprint of their effects on the test statistics of included variants, we reasoned that a model that uses the LD between omitted and tested variants should be able to assign that association to the omitted variant. To assess this, we developed an extension to SuSiE that explicitly models the probability that an omitted variant is causal, an approach that we call SuSiE-OUT. Rather than forcing every association signal to be explained by a tested variant, it allows each sparse effect to be assigned either to a tested variant or to an omitted variant; we call the second option an *OUT state* and its posterior inclusion probability an *OUT KIK*. Each candidate’s *LD fiegevpviet* is its vector of correlations with the tested variants, and SuSiE-OUT scores each candidate by how well its fingerprint matches the observed association pattern (fig. 1c,d). SuSiE-OUT never uses association statistics for omitted variants, whether measured or imputed. It also includes variance components for diffuse polygenic signal [41] and for LD mismatch [22], and it averages over a grid of their settings, weighted by fit [46] (Methods).

We compared SuSiE-OUT and masked-SuSiE with two alternatives. SuSiE-inf is fit to the tested variants and adds an infinitesimal component for many small effects [41]. Impute-then-SuSiE predicts association statistics for omitted variants from their LD with tested variants, keeps those with *q* ≥ 0.3 and fine-maps the combined set [36, 37]. Imputation and SuSiE-OUT therefore require the same input: LD between tested and omitted variants.

SuSiE-inf did not reliably correct filter-induced errors. With the SNP-only filter, it still made 40 false discoveries, compared with 52 for masked-SuSiE, and recalled only 63% of reference positives, compared with 97% (fig. 4a,b). With the MAf filter, it halved false discoveries, but only by making far fewer calls (fig. 4c). This is consistent with an omitted causal variant acting as a missing sparse effect, which a component for many small background effects is not designed to capture.

**Fig. 4.**
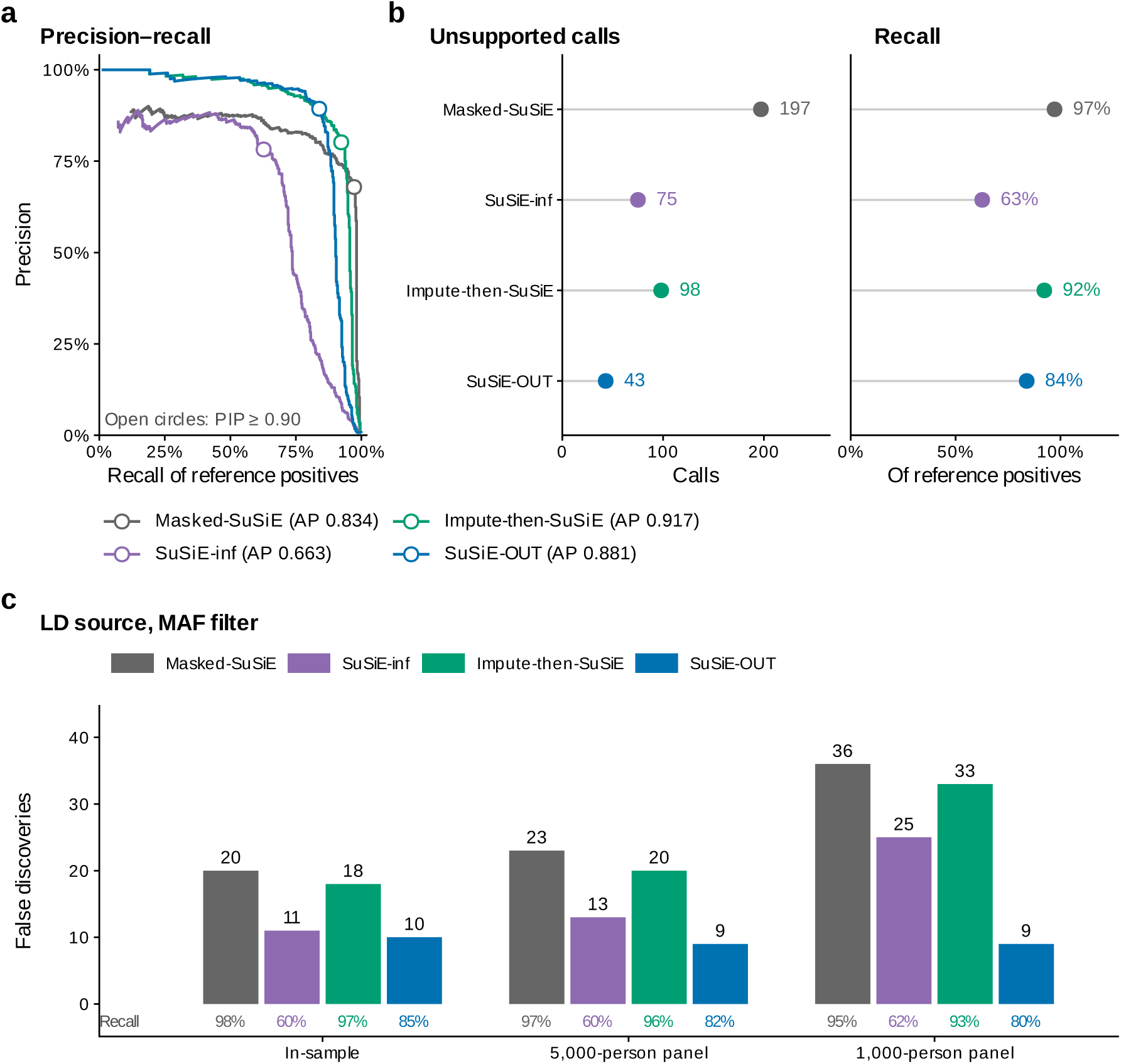
Imputing or modeling omitted variants reduces filter-induced false discoveries; SuSiE-inf does not reliably reduce them. All metrics measure agreement with unfiltered-SuSiE at 499 All of Us loci; a reference positive has reference PIP of at least 0.90. **a,** Precision–recall curves for tested variants under the SNP-only filter (429 reference positives); open circles indicate PIP 0.90. **b,** Unsupported calls (PIP 0.90, reference PIP *<* 0.90) and recall at PIP 0.90 under the same filter. **c,** false discoveries (PIP 0.90, reference PIP *<* 0.10) under the MAf filter, with LD among tested variants and between tested and omitted variants estimated in the analysis sample (in-sample) or in disjoint 5,000- or 1,000-person All of Us reference panels.

In contrast, with the SNP-only filter, imputation and SuSiE-OUT both substantially reduced false discoveries, with different trade-offs between recall and unsupported calls. Relative to masked-SuSiE, false discoveries fell from 52 to 17 with imputation and to 18 with SuSiE-OUT. Imputation recalled more reference positives (92% versus 84% for SuSiE-OUT) and had the highest overall agreement with unfiltered-SuSiE (average precision; fig. 4a). SuSiE-OUT made fewer unsupported calls (43, compared with 98 for imputation and 197 for masked-SuSiE; fig. 4b).

In practice, LD between variants often comes from a reference panel rather than from the GWAS genotype data [5, 10, 22]. We therefore repeated the MAf-filter analysis with LD from disjoint All of Us panels of 5,000 or 1,000 people. With panel LD, masked-SuSiE and impute-then-SuSiE made two to four times as many false discoveries as SuSiE-OUT (fig. 4c). However, SuSiE-OUT had lower recall (about 80% versus 93–97%).

### Omitting the blood-group O frameshift creates false discoveries at *ABO*

We observed that the *RBO* locus shows how filtering can mistakenly replace a well-characterized functional variant with confident but incorrect proxy calls (fig. 5). The single-base deletion rs8176719 defines blood group O. The frameshift truncates the ABO glycosyltransferase, so people with blood group O, who carry two copies, lack the active enzyme [47]. *RBO* is also an established LDL-C locus [48, 44]. Variants at *RBO* are associated with serum levels of plant sterols, which are markers of intestinal cholesterol absorption, so *RBO* likely affects LDL-C at least partly through cholesterol uptake in the gut [49]; blood group O is also associated with lower risk of myocardial infarction [50]. In the LDL-C analysis, rs8176719 had the strongest association in the region (*P* = 3 × 10^−22^), was tagged (*q* = 0.98) and had reference PIP 1.00. With indels omitted, masked-SuSiE instead gave PIPs above 0.999 to three linked SNPs whose reference PIPs were only 0.003–0.01. A workflow that acted on these calls would select three unsupported variants for functional follow-up and would not select the blood-group O allele. SuSiE-inf also gave all three SNPs PIP 1.00.

**Fig. 5.**
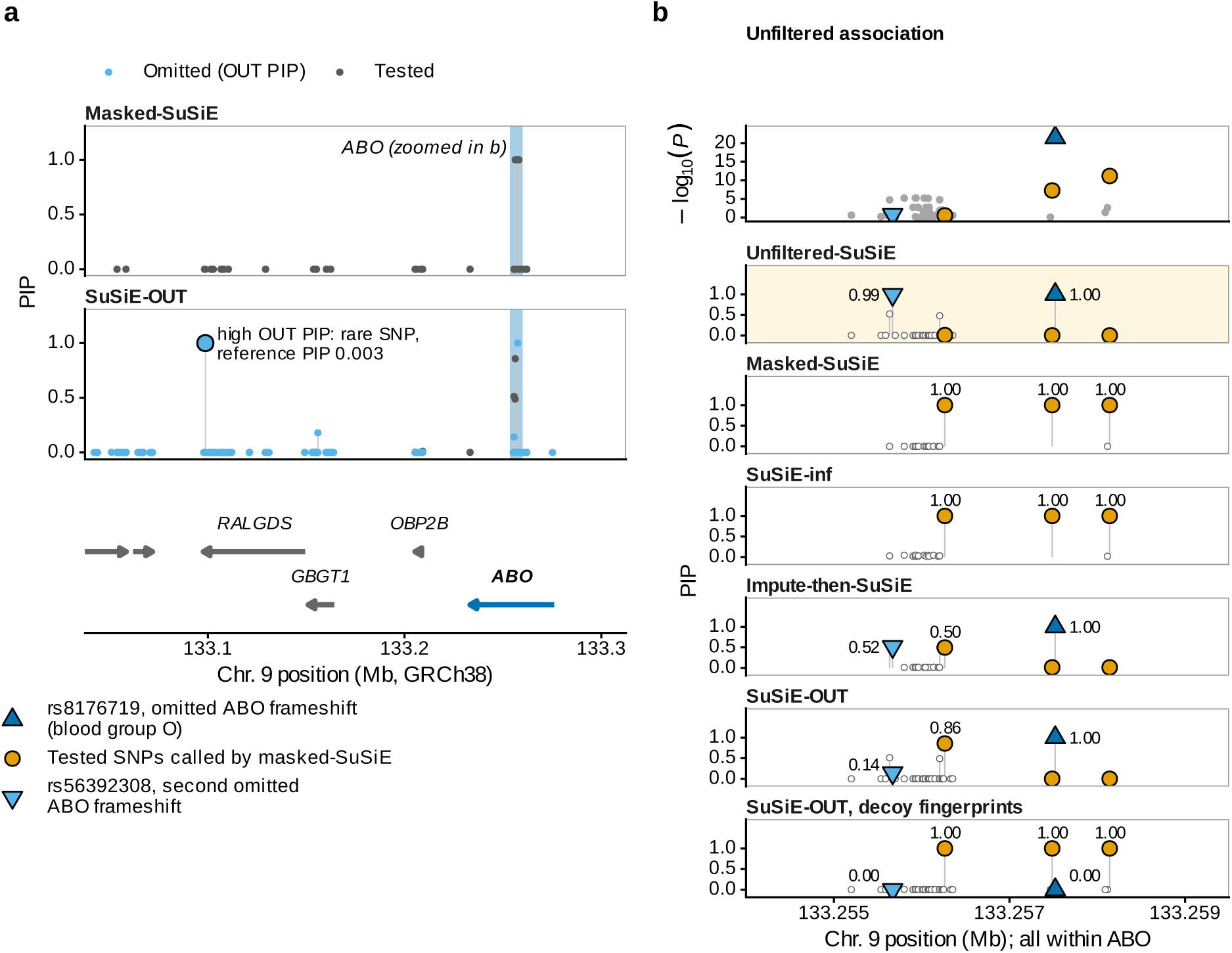
Omitting the *ABO* blood-group O frameshift creates false fine-mapping calls. LDL cholesterol in All of Us, SNP-only filter. The omitted indel rs8176719 (9:133257521:T:TC; tagging *q* = 0.98) is the frameshift that defines blood group O (GRCh38 carries the O allele). Other loci at which a masked-SuSiE false discovery coincided with a tagged omitted indel are listed in Supplementary Table 2. **a,** PIPs under masked-SuSiE and SuSiE-OUT for the 250 kb around *RBO*; blue points are omitted variants scored as OUT states. No variant elsewhere in the 1-Mb fine-mapping window had PIP above 0.05 under either method or unfiltered-SuSiE. SuSiE-OUT also gave OUT PIP 1.00 to a rare SNP in *RRLGDS*, 160 kb from rs8176719 (outlined; reference PIP 0.003). The gene track shows Ensembl [51] protein-coding genes; small genes are unlabeled. **b,** Within *RBO*: log_10_ *P* from the unfiltered association analysis (top) and PIPs for each method. Unfiltered-SuSiE (gold) analyzed all variants and gave PIP 1.00 to rs8176719 (blue triangle) and 0.99 to a second omitted *RBO* frameshift, rs56392308 (light-blue inverted triangle), which had little marginal association. With both indels omitted, masked-SuSiE and SuSiE-inf gave PIP 1.00 to three tested SNPs (orange) whose reference PIPs were 0.003–0.010. Impute-then-SuSiE and SuSiE-OUT each gave rs8176719 PIP 1.00 and lowered two of the three SNPs to 0.015 or less. The third SNP remained at 0.50 with imputation and 0.86 with SuSiE-OUT, and the second indel received 0.52 and 0.14. SuSiE-OUT with decoy fingerprints (circularly shifted across tested variants) gave all three SNPs PIP 1.00 and gave the frameshift PIP *<* 0.001. The correction therefore depends on the true LD to the omitted variant.

At *RBO*, impute-then-SuSiE and SuSiE-OUT both gave rs8176719 PIP 1.00 and lowered the PIPs of the two nearest proxies to 0.015 or less. The third proxy tagged a second omitted *RBO* frameshift, rs56392308. This indel had little marginal association but reference PIP 0.99. Both remedies divided posterior probability between this indel and its proxy. However, SuSiE-OUT also gave OUT PIP 1.00 to a distant rare SNP with reference PIP 0.003 (fig. 5a). This suggests that a high OUT PIP indicates association that the tested variants do not explain, not that a specific omitted variant is necessarily causal.

We observed the same pattern at *LCORL*, one of the strongest height loci [42]. Omitting an in-frame deletion gave two proxies near-certain PIPs. SuSiE-inf still gave one of them PIP 0.76, whereas both remedies lowered both below 0.02 (Supplementary fig. 1).

### SuSiE-OUT’s correction requires the true LD to omitted variants

We next asked whether SuSiE-OUT corrected false discoveries by matching the specific LD patterns of omitted variants, or merely by allowing association to be assigned outside the tested set. To distinguish these explanations, we refit SuSiE-OUT with *decoy* fingerprints. Decoys keep the same LD values but assign them to the wrong tested variants. We built decoys in three ways: by circularly shifting each fingerprint, by permuting the tested variants or by using fingerprints from another locus (*foveige* decoys).

In simulations, decoy fingerprints reduced false discoveries far less than true fingerprints did. In the main simulation, at loci with an omitted causal variant, SuSiE-OUT with true fingerprints removed 74% of masked-SuSiE false discoveries, whereas decoys removed only 7–8%. Across all six simulation settings, including rare omitted variants, hundreds of OUT candidates and polygenic background, true fingerprints removed 74–100% and decoys 4–58% (fig. 3b; Supplementary Table 3).

Our simulations at loci with tagged omitted variants recapitulated similar behavior in All of Us. At the 156 loci with a tagged omitted indel, true fingerprints removed 64% of masked-SuSiE false discoveries (42 to 15), whereas decoys removed 29–33% (to 28–30; fig. 3c). Among the 159 loci with an omitted credible-set indel, true fingerprints gave the highest OUT PIP to that indel at 37% (59/159); decoys almost never did (fig. 3d). Under the MAf filter, where omitted variants were mostly untagged, decoys did as well as true fingerprints (Supplementary Table 4). This is expected, because the fingerprints of untagged variants carry little information. Therefore, SuSiE-OUT’s correction depends on the true LD between omitted and tested variants; OUT states without that LD do not reproduce it.

### MAF thresholds create false discoveries in public UKB fine-mapping

Finally, we asked how often MAf thresholds would change calls in UKB fine-mapping analyses. We analyzed 169 loci across ten traits from a UKB SuSiE fine-mapping of 100,000 participants [41, 30]. At each MAf threshold, we refit masked-SuSiE and SuSiE-OUT to the variants that passed it and used the PIPs from the unfiltered analysis as a silver standard; here a false discovery is a call with an unfiltered PIP below 0.10. A 0.5%, 1% or 2% MAf threshold omitted a variant in the public credible set at 7%, 15% and 25% of loci, respectively (fig. 6a).

**Fig. 6.**
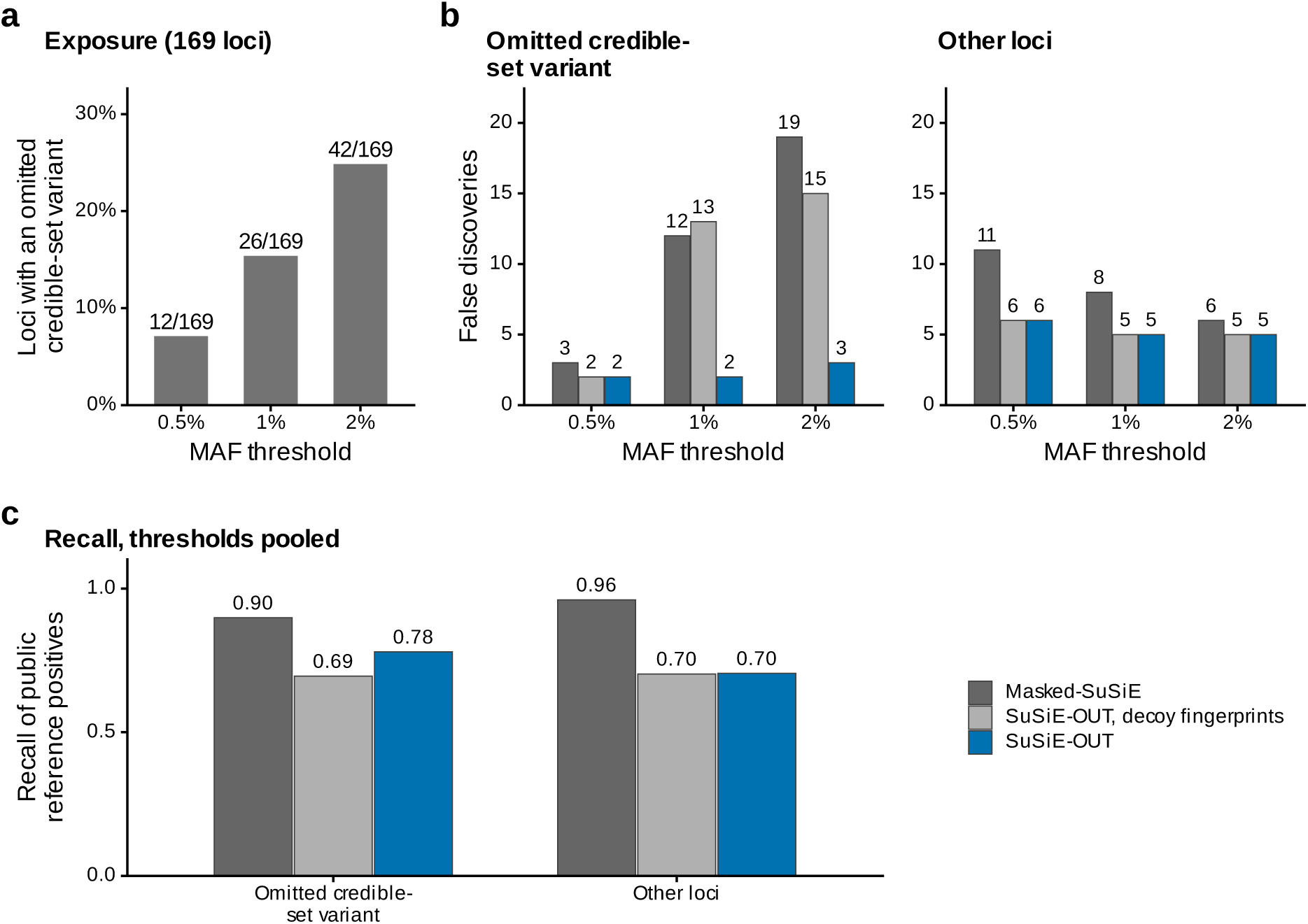
MAF thresholds create false discoveries in public UKB fine-mapping. 169 loci across ten traits from the public UK Biobank SuSiE fine-mapping of 100,000 participants. At each threshold, we refit masked-SuSiE and SuSiE-OUT to the variants passing it, using public UKB LD. The reference is the public PIP from the analysis of all variants, not causal truth. **a,** Loci whose public 95% credible set contains a variant omitted by the threshold. **b,** false discoveries (fitted PIP 0.90, public PIP *<* 0.10) at loci where the threshold omitted a credible-set variant (left) and at other loci (right). Gray, masked-SuSiE; light gray, SuSiE-OUT with decoy fingerprints (circularly shifted across tested variants or taken from a size-matched locus on another chromosome; mean of the two); blue, SuSiE-OUT. **c,** Recall of public reference positives (public PIP 0.90), pooled over the three thresholds, by locus type. At loci without an omitted credible-set variant, SuSiE-OUT with true and with decoy fingerprints had nearly the same recall. The lower recall there therefore comes from the LD-mismatch and polygenic components rather than from the OUT states. Results at a 5% threshold are in Supplementary Table 5.

At loci where the MAf filtering omitted a credible-set variant, masked-SuSiE made 3, 12 and 19 false discoveries at the 0.5%, 1% and 2% thresholds compared to SuSiE-OUT, which made 2, 2 and 3 false discoveries. In contrast, SuSiE-OUT with decoy fingerprints made 2, 13 and 15, nearly as many as masked-SuSiE (fig. 6b).

SuSiE-OUT’s reduction in false discoveries came with lower recall. At loci where the threshold omitted no credible-set variant, SuSiE-OUT had lower recall than masked-SuSiE (71% versus 96%), and recall was nearly the same with true and decoy fingerprints. This suggests that the lower recall comes from SuSiE-OUT’s polygenic and LD-mismatch components rather than from its OUT states (Supplementary Table 5). Across all 169 loci at the 1% threshold, SuSiE-OUT made 13 fewer false discoveries than masked-SuSiE (7 versus 20) and 47 fewer supported calls (134 versus 181). SuSiE-OUT is therefore preferable for users who consider a false discovery at least 3.6 times as costly as a missed supported call; this break-even ratio was 8.2 at the 0.5% threshold and 2.4 at the 2% threshold.

## Discussion

Human genetics concerns the discovery of causal genetic variants; thus, fine-mapping procedures represent an essential component of modern human genetics [4, 21]. Although fine-mapping works remarkably well in simulations [8, 9, 22], in practice fine-mapping algorithms are applied with choices that materially affect the underlying method assumptions [5, 52]. Most importantly, fine-mapping distributes posterior probability only among the tested variants. It does not account for variants that were omitted before fine-mapping. Two common choices in fine-mapping workflows are a 1% MAf threshold and the exclusion of indels, and both clearly can omit causal variants. In All of Us, these filters resulted in 20–32% of calls that were discrepant with an unfiltered fine-mapping approach. fine-mapping studies should therefore be highly explicit about variant filtering and QC procedures that may inadvertently result in uninterpretable credible sets.

Furthermore, these kinds of variant omissions are enriched in variants that are deleterious and therefore more likely to be causal. The filters preferentially omit rare, larger-effect alleles and coding indels, which often truncate or alter protein sequence and are strongly depleted from constrained genes [26]. fine-mapping then assigns their association to linked SNPs with near-certain PIPs. At *RBO*, three linked SNPs received the high PIPs that unfiltered-SuSiE gave to the blood-group O allele and a second frameshift. At *LCORL*, two linked SNPs received the high PIP that unfiltered-SuSiE gave to an in-frame deletion. Researchers use high-PIP variants to assign genes, to choose variants for reporter assays and genome editing, and to build sets of putatively causal variants for studying regulatory mechanisms [15, 16, 17, 18, 19, 20, 21]. filtering affects each of these use cases. The same errors should arise from any filter that omits tagged causal variants, including imputation-quality filters and restriction to variants present in an external LD reference; restricting finnGen fine-mapping to variants in a 1000 Genomes reference produced such calls, and SuSiE-OUT lowered 15 of 18 below PIP 0.90 (Extended Data fig. 1).

We present two remedies that recover some of these omitted causal variant effects: SuSiE-OUT and impute-then-SuSiE. Both require LD between tested and omitted variants, which fine-mapping releases rarely provide; releasing the omitted variants and their LD with tested variants would let others apply either remedy. Both remedies reduced errors, and each performed better in a different setting. With in-sample LD, imputation had higher recall and agreed best with unfiltered-SuSiE. In contrast, with LD from a small external panel, as in most public fine-mapping, SuSiE-OUT made far fewer false discoveries but had lower recall (fig. 4c). SuSiE-OUT also does not require imputed association statistics. It gave the highest OUT PIP to the omitted indel at more than a third of loci with an omitted credible-set indel. However, it sometimes gave high OUT PIPs to variants that unfiltered-SuSiE did not support. OUT PIPs should therefore be read as evidence that some causal variation is missing from the tested set, not as calls. SuSiE-inf, which models diffuse background effects [41], reduced errors only by making fewer calls. This is consistent with an omitted causal variant acting as a missing sparse effect.

Our study has three limitations. first, the silver standard in All of Us and UKB is unfiltered fine-mapping, not a truly orthogonal causal truth. Second, although SuSiE-OUT was in aggregate beneficial at loci with omitted causal variants, SuSiE-OUT’s probability that at least one causal variant was omitted (*locus OUT suppovt*) was somewhat unreliable at highly polygenic loci (Supplementary Note 1; Extended Data figs. 2 and 3). finally, our All of Us reference used the exome callset, so it omits most noncoding variation, which harbors most trait-associated variants [45, 18]. Our estimates are therefore likely conservative. When we refit only the unfiltered reference at 25 loci using genome-wide rather than exome sequence, several exome variants with near-certain reference PIPs appeared to be proxies for noncoding variants with stronger associations, and most calls at these loci were no longer supported (Supplementary Note 2).

Overall, we describe highly prevalent failure modes in fine-mapping that occur through mundane QC and variant filtering procedures. These procedures miscalibrate standard fine-mapping PIPs since these methods assume that all causal variants have been included; miscalibration of fine-mapping in real data has also been reported from other sources [52, 41]. Moving forward, we recommend that fine-mapping results from “standard” post-GWAS workflows (which typically filter low-frequency variants at some threshold) use slightly more cautious framing than “causal” variants or “credible sets”, since these labels imply slightly greater confidence than generally is warranted; we recommend “putatively causal” or “fine-mapping prioritized variants”. We emphasize that despite elevated false discovery rates of standard fine-mapping approaches, such approaches are likely still considerably superior to using marginal GWAS summary statistics for nominating causal variants [6, 4]. We suggest either using weaker claims of causality, or the application of fine-mapping procedures (e.g., SuSiE-OUT or impute-then-SuSiE) that are better calibrated in real-world scenarios.

## Online Methods

### Terms

*Tested* variants pass a filter and enter fine-mapping; *omitted* variants are excluded by it. *Masked-SuSiE* is ordinary SuSiE fit to tested variants. *Uefilteved-SuSiE* is SuSiE fit to tested and omitted variants together and is the reference in All of Us; in UKB the reference is the public PIP from the published analysis of all variants. Calls, unsupported calls and false discoveries are defined in Table 1. All SuSiE, SuSiE-OUT and unfiltered fits used at most *L* = 10 sparse effects, and all SuSiE-OUT fits used SuSiEOut version 0.2.0, an R package interoperable with susieR [9], with prior weights estimated once on simulated development loci and then frozen. Implementation details and software tests are in the Supplementary Methods.

### SuSiE-OUT statistical model

SuSiE-OUT extends the single-effect decomposition used by SuSiE [9, 22]. for sparse single-effect component *l*, the model assigns one unobserved state:

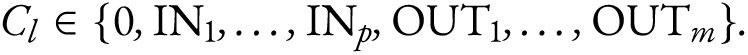

Here, the *p* tested variants have GWAS statistics, the *m* omitted variants are present only in a sequencing reference panel, and state 0 means no sparse effect. Let *z_S_* be the vector of *z*-statistics for tested variants, *f_k_* variant *k*’s LD fingerprint (its correlations with all tested variants), *n* the GWAS sample size and Σ_0_ the residual covariance. Given standardized effect *β*,

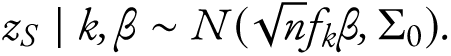

for normal-mixture component *t* we integrated *β* ∼ *N* (*μ_t_, v_t_*). Let *r_l_* be the residual *z*-statistic vector for component 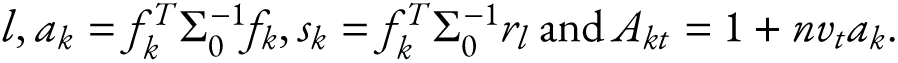 The rank-one log Bayes factor against no sparse effect is

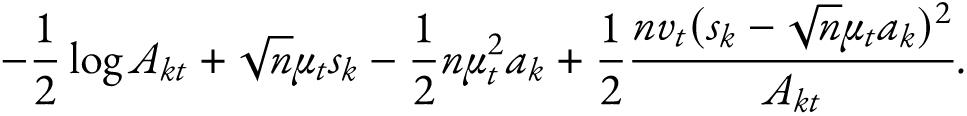

Attribution therefore depends on the full fingerprint, not on a separate marginal-effect estimate. Iterative Bayesian stepwise updates [9] revised each component’s residuals and posterior weights for the 0, IN and OUT states. Within covariance setting *g*, with 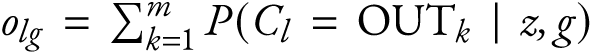 the OUT probability of component *l*, locus OUT support is 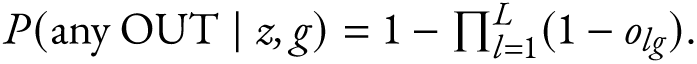.

### Dense and LD-mismatch components

The model covariance was

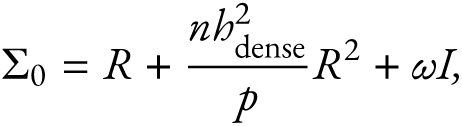

where *R* is the analysis LD matrix and *R*^2^ = *RR*. The dense term adds variance along directions with stronger LD, as expected when many correlated variants have small effects; the isotropic term *ω* captures general disagreement between the association pattern and the reference LD. The model was fit under each of 12 settings, *b*^2^_dense_ ∈ {0, 0.002, 0.008, 0.032} and *ω* ∈ {0, 0.04, 0.12}. With *R* = *U* diag(*λ_i_*)*U^T^*, the conditional fit supplied posterior mean residual *r̃_i_* and sparse posterior uncertainty *v_i_* in eigendirection *i*, and the expected Gaussian negative log-likelihood for setting *g* = (*h*^2^*, ω*) was

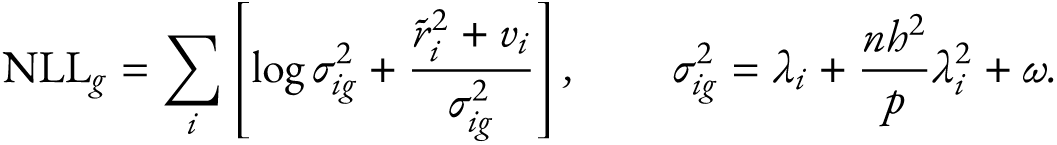

Prior weights *π_g_* for the settings, and a prior on the number of sparse effects, were estimated by label-free finite-mixture expectation-maximization (EM [53]; Dirichlet strength 5, at most 50 updates) on an independent development set of simulated loci. Posterior setting weights were *γ_g_* ∝ *π_g_* exp(− NLL*_g_*/2), and PIPs and locus OUT support were averaged over settings with these weights. The LD-mismatch posterior is Σ*_g_*_:*ωg>*0_ *γ_g_*. All analyses applied the development-set weights unchanged, and no analysis used omitted-variant association statistics to fit or select a model.

## Simulations

### Main simulation

We generated 600 evaluation loci with 40 tested and 12 omitted variants using the package’s Gaussian locus simulator, with first-order autoregressive LD (adjacent-variant correlation 0.88). Each omitted variant was assigned one of five exclusion mechanisms in rotation: MAf below 1%, indel, imputation quality below 0.8, absence from the LD reference, or monomorphism in the finite reference panel. Loci had *coetaieed*, *pavtial-omissioe* or *all-omitted* sparse causes, in which the tested set included all, some or none of the one to six causal variants, or were null, polygenic (local *h*^2^ = 0.008) or LD-mismatch (isotropic variance 0.08) controls. Three settings were distributed across loci: neutral selection with GWAS *n* = 100,000 and a 500-sample reference panel; stabilizing selection with *n* = 500,000 and a 1,000-sample panel; and directional selection with *n* = 1,000,000 and exact population LD. Under neutral selection, effects were drawn from *N* (0, 0.018^2^); under stabilizing selection, the effect standard deviation was 0.012 (MAf/0.01)^−0.18^; under directional selection, effects had mean 0.018 and signs aligned with the derived allele. Within each setting, half of the loci used a reference panel from a population with weaker LD (adjacent-variant correlation 0.72). Priors were estimated on a separate development set of 1,800 loci without architecture labels.

The 600 loci comprised 292 contained, 118 partial-omission, 134 all-omitted, 18 null, 19 polygenic and 19 LD-mismatch loci (Supplementary Table 1). A false discovery was a noncausal tested variant with PIP ≥ 0.90, and power was the proportion of contained loci with a causal tested variant at PIP ≥ 0.90. Tagging of an omitted causal variant was the maximum *r*^2^ in the analysis population between any omitted causal variant and any tested variant. Intervals used 2,000 bootstrap resamples of loci within strata, and locus-level comparisons of false-discovery occurrence used an exact McNemar test.

### Feature-by-feature simulations

A second simulator added realistic features one at a time. Each locus had 40 tested variants at uniform positions with latent correlation exp(−*d*/0.15), one to three tested causal variants (noncentrality 6–12, random sign) and, with probability 0.42, one or two omitted causal variants (noncentrality 6–15); *n* = 500,000. The baseline placed 12 common omitted variants at one end of the locus. Added features were omitted variants interleaved with tested ones; genotype-based LD from 50,000 simulated individuals with omitted variants of MAf 0.05–1% (rare omitted variants); 400 rather than 12 OUT candidates; extra tested signals; small omitted effects; LD from a 500-person reference with weaker LD; and polygenic background (noncentrality SD 0.3 or 1.0 added to every variant). Omitted variants without LD to any tested variant were not OUT candidates. We simulated 500 loci per condition. Dense-grid refits used for Supplementary Note 1 are described in the Supplementary Methods.

### Decoy-fingerprint simulations

To test whether SuSiE-OUT’s correction depends on the true LD between tested and omitted variants, we refit it with decoy fingerprints that keep the values of the tested-by-omitted LD matrix *C* but break its alignment with the tested variants. The *civculav-shift* decoy shifted each column of *C* by an independent nonzero offset across tested variants; the *pevmutatioe* decoy applied one random permutation of tested variants to all columns; and the *foveige* decoy took columns from another simulated locus of the same condition, resampled to the recipient’s number of omitted variants. foreign decoys were used only with the feature-by-feature simulator, because all main-simulation loci share the same autoregressive LD structure. We applied the decoys to all 600 main-simulation loci (five realizations each) and to 500 loci in each of five feature-by-feature conditions (three realizations each), with loci and seeds identical to the original analyses. Every arm, together with masked-SuSiE and unfiltered-SuSiE, was scored against truth and, separately, against unfiltered-SuSiE. Decoy arms were averaged over realizations within a locus, and decoy-minus-true differences used 2,000 locus-bootstrap resamples (Supplementary Table 3). The true-LD arm reproduced the original benchmark exactly.

### All of Us analyses

#### Participants, phenotypes and loci

We used All of Us [38, 39] Controlled Tier release C2025Q4R6 (version 9) and its short-read whole-genome-sequencing exome PGEN callset (GRCh38), restricted to active, unrelated participants of the European-ancestry group who passed sample filters and had complete phenotype and covariate data. Height (cm), LDL-C (mg dl^-1^; divided by 0.70 during lipid-lowering treatment) and platelet count (10^3^ cells μl^-1^) were range-checked and summarized by the participant median. Each phenotype was regressed on age, age squared, sex, age-by-sex and ten genetic principal components [54], and residuals were transformed to normal scores. Sample sizes were 275,875 for height, 137,377 for LDL-C and 141,410 for platelet count.

Loci were selected per phenotype from autosomal biallelic variants with MAf of at least 1%, call rate of at least 99% and Hardy–Weinberg *P >* 10^−6^: genome-wide-significant leads (*P <* 5 × 10^−8^) were retained greedily if more than 1 Mb from a retained lead, with regions of ±500 kb. This yielded 367 height, 31 LDL-C and 101 platelet-count loci. Local variants passed the release filter, call rate of at least 99%, minor allele count of at least 20 and Hardy–Weinberg *P >* 10^−6^. Additive association statistics were computed with PLINK 2 [55], and signed genotype correlations in the same participants; LD matrix checks and repair are described in the Supplementary Methods.

#### Filtering designs

In the *MRF filtev* design, tested variants had MAf of at least 1% and omitted variants had MAf below 1%. In the *SMK-oely filtev* design, the callset was rebuilt so that indels could be analyzed: multiallelic records were split with PLINK 2 and left-normalized against GRCh38 before the same quality control. Tested variants were SNPs with MAf of at least 1%, and every other quality-controlled variant, including every indel, was omitted: 60,218 tested and 617,045 omitted variants, of which 52,295 were indels (Supplementary Table 6). Each design was compared with its own unfiltered-SuSiE fit to all its variants (prior variance estimated, residual variance fixed, at most 1,000 iterations). Because the callsets differ, the designs were not compared head to head. We confirmed at all 499 loci that tested LD and *C* equalled the corresponding blocks of the unfiltered LD matrix and that identifiers, alleles, row order, *z*-statistics and reference bases agreed across files.

#### Methods compared

Masked-SuSiE used the settings of unfiltered-SuSiE. SuSiE-inf used the authors’ implementation [41] with method-of-moments variance estimation and at most 1,000 iterations; when moments estimation did not converge, the maximum-likelihood estimator was used (two loci in the MAf-filter design and four in the SNP-only design). SuSiE-OUT used tested-variant statistics, tested LD and *C*, with the frozen priors, the three most probable covariance settings (at least 95% of the prior mass), tolerance 10^−5^ and at most 1,000 iterations. No method other than unfiltered-SuSiE used measured omitted-variant statistics.

#### Imputation and tagging

For impute-then-SuSiE [36, 37], with tested-variant statistics *z_S_*, tested-variant LD matrix *R*, omitted-by-tested LD matrix *C* and a fixed ridge *λ* = 10^−4^, we computed *W* = *C* (*R* + *λI*)^−1^ and imputation quality *q_j_* = (*WRW* ^T^)*_jj_*. Eligible rows (*q_j_* ≥ 0.3) were normalized as 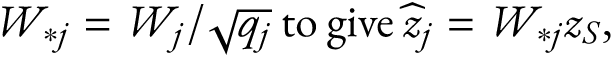 and SuSiE was fit to observed and predicted statistics with covariance

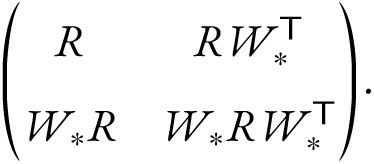

Measured omitted-variant statistics were used only after fitting, to evaluate prediction (Supplementary Table 7 reports *q >* 0 and *q* ≥ 0.8 thresholds).

We used the same LD to measure tagging, the fraction of an omitted variant’s signal predictable from tested variants: 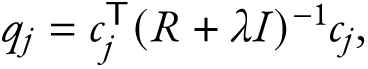 with *c_j_* its fingerprint, which equals the imputation quality up to the ridge term. Predicted statistics 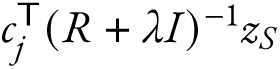 agreed with measured statistics for omitted indels with *q* ≥ 0.8 (correlation 0.998, slope 1.00); decoy fingerprints gave correlations near zero.

An *omissioe eveet* was an unfiltered-SuSiE 95% credible set (after the purity filter [9]) containing an omitted variant. In the SNP-only design, loci were *iedel eveets* if such a set contained an omitted indel, *vave-SMK-oely eveets* if it contained only omitted SNPs, and otherwise had no event; events were stratified by the largest *q* among omitted credible-set variants (*<* 0.1, 0.1 to *<* 0.3, ≥ 0.3).

#### Evaluation

Terms followed Table 1, with unfiltered-SuSiE as the reference and tested variants as candidates. Precision was the fraction of calls that were supported, and average precision the stepwise area under the precision–recall curve over PIP thresholds, with tied PIPs entering together (536 reference positives in the MAf-filter design and 429 in the SNP-only design). Intervals for method differences used 2,000 bootstrap resamples of loci [56], with methods paired within loci (Supplementary Table 8).

#### Decoy fingerprints in All of Us

In the SNP-only design, SuSiE-OUT was refit at all 499 loci with circular-shift decoys and with foreign decoys from the locus on another chromosome with the closest numbers of tested and omitted variants, resampled to the recipient’s dimensions. Each decoy had three seeded realizations, averaged within loci; decoy-minus-true differences used 2,000 locus-bootstrap resamples. The MAf-filter design used one realization of each (Supplementary Table 4). A locus was *ideetified* when the omitted variant with the highest SuSiE-OUT PIP belonged to an unfiltered-SuSiE credible set.

#### Example loci

In the SNP-only design, we listed loci at which an omitted indel with *q* ≥ 0.3 and unfiltered-SuSiE PIP of at least 0.5 coincided with a masked-SuSiE false discovery, ranked by the indel’s reference PIP and then its absolute *z*. Twenty-four loci (40 indel–proxy pairs) qualified (Supplementary Table 2). At *RBO* (fig. 5), the omitted indel rs8176719 is the single-base frameshift that defines blood group O; GRCh38 carries the O deletion, so the indel is represented as an insertion of the non-O allele. Gene positions in fig. 5 are from Ensembl (GRCh38).

#### External reference LD

We drew nested samples of 1,000 and 5,000 European-ancestry All of Us participants who passed quality control, excluding related participants and everyone in the association analyses. In the MAf-filter design, tested LD and *C* were recomputed in each panel, with association statistics unchanged, and masked-SuSiE, SuSiE-inf, impute-then-SuSiE and SuSiE-OUT were refit. SuSiE-inf with panel LD set the infinitesimal variance to zero when estimation failed (8 loci with the 1,000-person panel and 4 with the 5,000-person panel; flagged in the source data). further details are in the Supplementary Methods.

#### UKB MAF-threshold analysis

We fixed the UKB inputs to Zenodo record 7055906 [41, 30]; UK Biobank is described elsewhere [40]. from the maximum-PIP lead of each trait-region across ten traits, 169 loci had a SNP lead and a compatible public Broad/Alkes UKB LD matrix [10] after liftover to GRCh38 and allele matching. The reference was the public SuSiE SNP result. At MAf thresholds of 0.5%, 1%, 2% and 5%, variants passing the threshold formed the tested set and matched variants with MAf between 0.1% and the threshold formed the omitted set; the public matrix supplied tested LD and *C*, and omitted-variant statistics were not inputs. Masked-SuSiE and SuSiE-OUT (at most 200 iterations, tolerance 10^−5^) were refit at every threshold. A locus was *affected* if an omitted variant was in its public credible set; every such variant had *q* ≥ 0.1. fig. 6b reports differences in counts, not paired changes, and fig. 6c pools recall over the 0.5%, 1% and 2% thresholds.

For the decoy analysis, SuSiE-OUT was refit at every locus and threshold with a circular-shift decoy and a size-matched foreign decoy from another chromosome. Saved real-fingerprint and masked-SuSiE fits were reused; refitting every 25th contrast reproduced them (maximum PIP difference 5 × 10^−16^). Decoy-minus-true differences used 1,000 bootstrap resamples of loci (Supplementary Table 5). Comparisons of public LD sources and sensitivity analyses are in Supplementary Tables 10 and 11 and the Supplementary Methods.

#### FinnGen external-reference filtering

To model an analysis without in-sample LD, we used finnGen release 12 summary statistics [57] with genotypes of 633 European 1000 Genomes participants [58, 28]. The tested set contained SNPs with finnGen MAf of at least 1% that were present in a prefiltered SNP-only 1000 Genomes panel (1000 Genomes MAf above 0.5%); the omitted set contained other variants, including indels, from the same participants’ sequence data with finnGen MAf of at least 0.05%. We fit 36 selected loci in 28 LD blocks and resampled blocks 2,000 times; finnGen PIPs were a benchmark, not causal truth (Supplementary Table 12; Extended Data fig. 1).

## Extended Data figure legends

**Extended Data Fig. 1.**
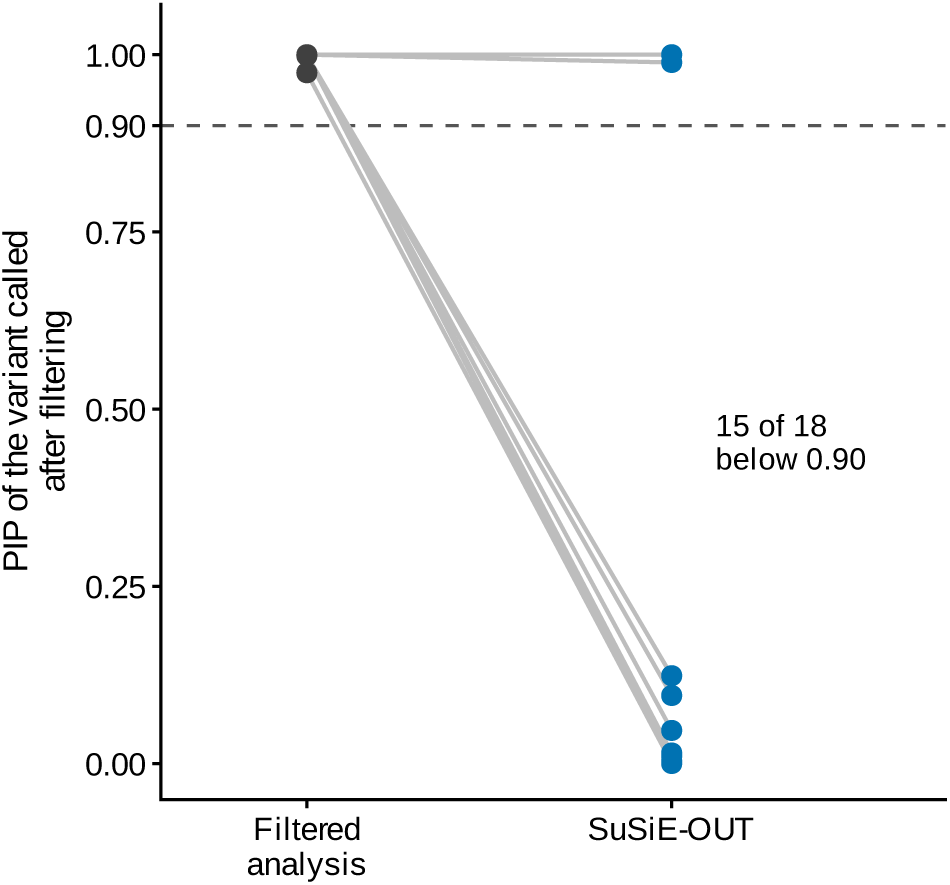
FinnGen external-reference panel. Each line represents one of 18 loci at which filtering to variants present in a 633-participant European-ancestry 1000 Genomes reference gave PIP of at least 0.90 to a variant other than the published finnGen lead; points compare the filtered-analysis and SuSiE-OUT PIPs. SuSiE-OUT lowered the PIP below 0.90 for 15 of these 18 variants. We selected the 36 loci to test a difficult case. Because the reference is small and ancestry-mismatched, these loci cannot estimate how often such errors occur or calibrate locus OUT support.

**Extended Data Fig. 2.**
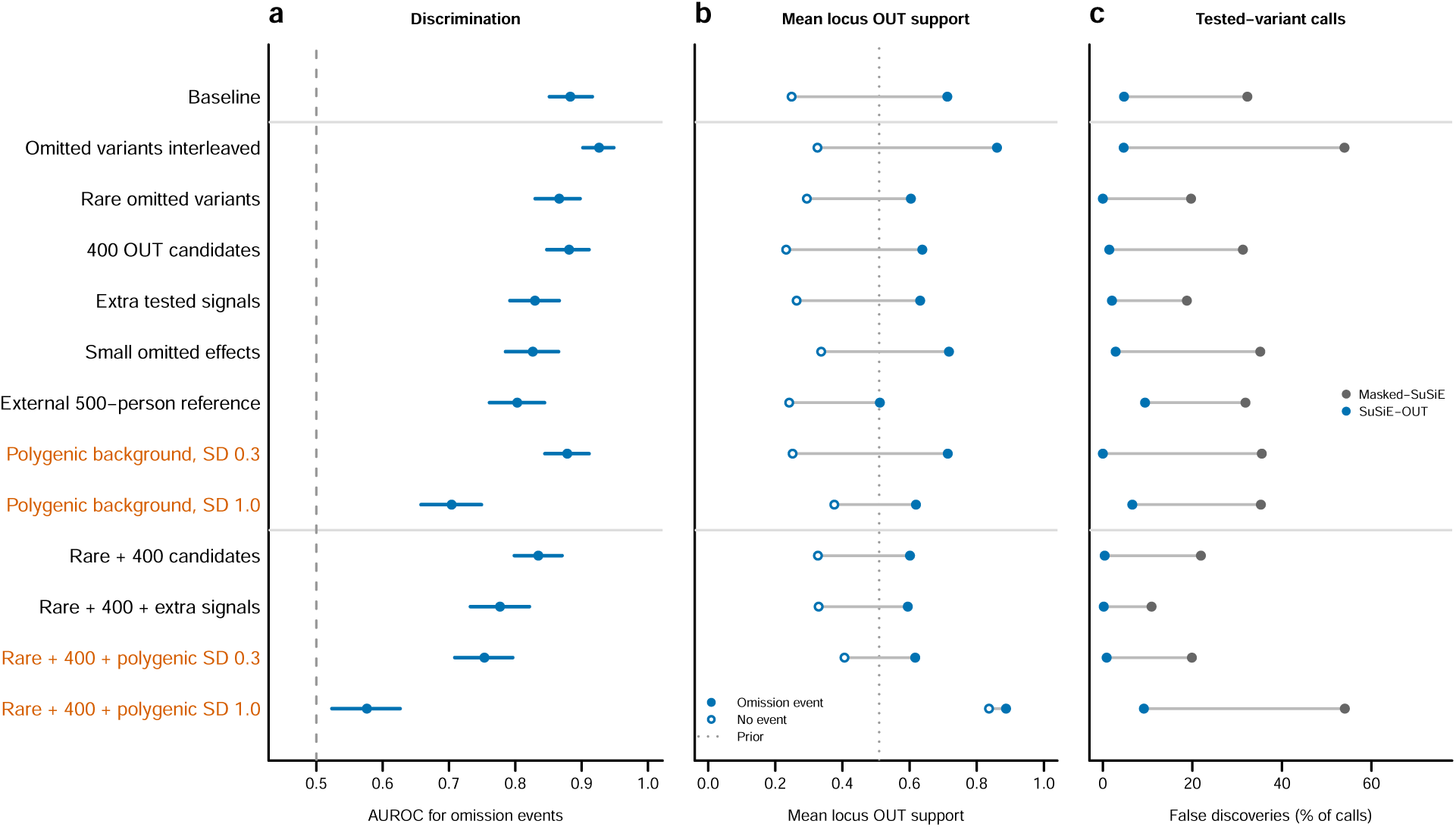
Polygenic background miscalibrates locus OUT support. Each row is a simulation condition with 500 loci, of which about 42% carried one or two large-effect omitted causal variants (omission events). The baseline mirrors the main simulations; the rows below it add one feature each, and the bottom four rows combine rare omitted variants and 400 OUT candidates, alone or with a third feature. Polygenic conditions are shown in orange; polygenic SD is the standard deviation of the background noncentrality added to every variant. **a,** AUROC of locus OUT support for omission events; bars are 95% locus-bootstrap intervals. **b,** Mean locus OUT support at event (filled) and non-event (open) loci; the dotted line is the prior probability of any OUT effect (0.51 by the Poisson approximation; 0.49 exactly at *L* = 10). **c,** false discoveries as a percentage of calls at event loci for masked-SuSiE (gray) and SuSiE-OUT (blue). All SuSiE-OUT fits used the fixed prior weights of the main simulations.

**Extended Data Fig. 3.**
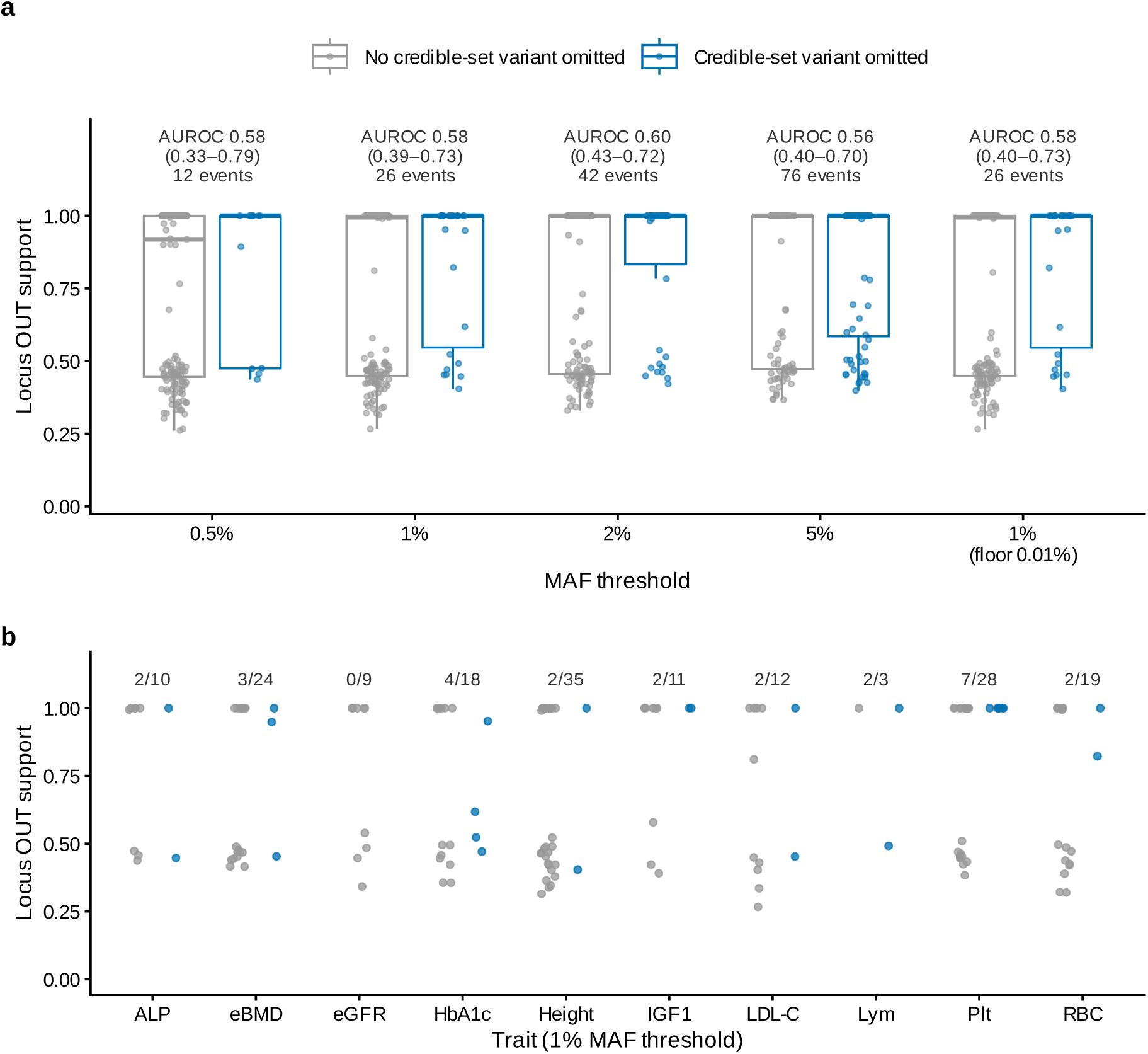
Locus OUT support in the UKB MAF-threshold analysis. a, Locus OUT support at the 169 UKB loci, by MAf threshold, for loci whose public credible set contained no variant omitted by the threshold (gray) or at least one (blue). Points are loci; boxes show medians and interquartile ranges. Labels give the AUROC of locus OUT support for an omitted credible-set variant, its 95% clustered-bootstrap interval (resampling traits and LD blocks, 2,000 replicates) and the number of such loci. The rightmost group keeps the 1% threshold but extends omitted variants to MAf 0.01%. **b,** Locus OUT support by trait at the 1% threshold; labels give loci with an omitted credible-set variant over all loci for that trait. Locus OUT support was close to 1 at many loci with and without such a variant, so it did not reliably identify incomplete tested sets. ALP, alkaline phosphatase; eBMD, estimated bone mineral density; eGfR, estimated glomerular filtration rate; HbA1c, hemoglobin A1c; IGf1, insulin-like growth factor 1; LDL-C, low-density lipoprotein cholesterol; Lym, lymphocyte count; Plt, platelet count; RBC, red blood cell count.

## Data availability

The analyses used released finnGen summary statistics, 1000 Genomes Project reference genotypes, public UK Biobank fine-mapping products and All of Us Controlled Tier data. Public-data manifests in the accompanying repository preserve the source URLs, release identifiers and checksums used in this study. All of Us individual-level data are available to authorized researchers through the Researcher Workbench.

## Code availability

The SuSiEOut R package is available at https://github.com/weinstocklab/SuSiEOut.

## Data Availability

This study used data from the All of Us Research Program's Controlled Tier Dataset CDRv9 available to authorized users on the Researcher Workbench.

## Acknowledgements

We thank members of the Weinstock lab for helpful feedback. We gratefully acknowledge *Rll of Us* participants for their contributions, without whom this research would not have been possible. We also thank the National Institutes of Health’s *Rll of Us* Research Program for making available the participant data [and/or samples and/or cohort] examined in this study.

## Correspondence

Correspondence should be addressed to Josh Weinstock. Opus 5.5 and gpt-6.1 were used to assist manuscript drafting, editing, and writing code.

